# Machine Learning Based Reanalysis of Clinical Scores for Distinguishing Between Ischemic and Hemorrhagic Stroke in Low Resource Setting

**DOI:** 10.1101/2022.03.03.22271885

**Authors:** Aman Bhardwaj, MV Padma Srivastava, Pulikottil Wilson Vinny, Amit Mehndiratta, Venugopalan Y Vishnu, Rahul Garg

## Abstract

**BACKGROUND:** Identification of stroke and classifying them as ischemic and hemorrhagic type using clinical scores alone faces two unaddressed issues. One pertains to over-estimation of performance of scores and the other involves class imbalance nature of stroke data leading to biased accuracy. We conducted a quantitative comparison of existing scores, after correcting them for the above-stated issues. We explored the utility of Machine Learning theory to address overestimation of performance and class imbalance inherent in these clinical scores.

**METHODS:** We included validation studies of Siriraj (SS), Guys Hospital/Allen (GHS/AS), Greek (GS), and Besson (BS) Scores for stroke classification, from 2001-2021, identified from systematic search on PubMed, ERIC, ScienceDirect, and IEEE-Xplore. From included studies we extracted the reported cross tabulation to identify the listed issues. Further, we mitigated them while recalculating all the performance metrics for a comparative analysis of the performance of SS, GHS/AS, GS, and BS.

**RESULTS:** A total of 21 studies were included. Our calculated sensitivity range (IS-diagnosis) for SS is 40-90% (median 70%[IQR:57-73%], aggregate 71%[SD:15%]) as against reported 43-97% (78%[IQR:65-88%]), for GHS/AS 35-93% (64%[IQR:53-71%], 64%[SD:17%]) against 35-94% (73%[IQR:62-88%]), and for GS 60-74% (64%[IQR:62-69%], 69%[SD:7%]) against 74-94% (89%[IQR:81-92%]). Calculated sensitivity (HS-diagnosis), for SS, GHS/AS, and GS respectively, are 34-86% (59%[IQR:50-79%], 61%[SD:17%]), 20-73% (46%[IQR:34-64%], 44%[SD:17%]), and 11-80% (43%[IQR:27-62%], 51%[SD:35%]) against reported 50-95% (71%[IQR:64-82%]), 33-93% (63%[IQR:39-73%]), and 41-80% (78%[IQR:59-79%]). Calculated accuracy ranges, are 37-86% (67%[IQR:56-75%], 68%[SD:13%]), 40-87% (58%[IQR:47-61%], 59%[SD:14%]), and 38-76% (51%[IQR:45-63%], 61%[SD:19%]) while the weighted accuracy ranges are 37-85% (64%[IQR:54-73%], 66%[SD:12%]), 43-80% (53%[IQR:47-62%], 54%[SD:13%]), and 38-77% (51%[IQR:44-64%], 60%[SD:20%]). Only one study evaluated BS.

**CONCLUSION:** Quantitative comparison of existing scores indicated significantly lower ranges of performance metrics as compared to the ones reported by the studies. We conclude that published clinical scores for stroke classification over-estimate performance. We recommend inclusion of equivocal predictions while calculating performance metrics for such analysis. Further, the high variability in performance of clinical scores in stroke identification and classification could be improved upon by creating a global data-pool with statistically important attributes. Scores based on Machine Learning from such globally pooled data may perform better and generalise at scale.

## Introduction

According to the Global Burden of Disease (GBD) analysis of 2019, for the population group of 50 years and above, stroke was the second major contributor to Disability Adjusted Life Years (DALYs), and the third leading cause of DALYs among the population of all age groups [1]. DALY is a measure of time and consists of the years of life lost due to early death and years of life spent with disability.

GBD analysis of 2016 reports that there were 5.5 million deaths globally due to stroke (2.9 million males, 2.6 million females) [2]. Global mortality due to stroke has shown a significant decline of 36.2% from 1990 to 2016 due to the advances in medical science, healthcare facilities and technology. Nations with high and high-middle Socio-Demographic Index (SDI) witnessed a larger decline in mortality of 51.9% and 44.7% respectively, compared to the nations with middle, low-middle, and low SDI with 38.2%, 22.7%, 20.8% declines respectively [2]. DALY follows a similar trend.

Developing countries bear burden of stroke more than the developed countries where 75.2% of total deaths and 81% of total DALYs are because of stroke [3]. With aging population, the developing countries may soon find themselves under an unbearable burden of stroke [4].

Ischemic Stroke (IS) and Hemorrhagic Stroke (HS), the two broad categories of stroke is heavily skewed towards the former with 84% prevalence for Ischemic type, however HS caused more fatalities than IS in 2016 (2.8 million and 2.7 million respectively) [2].

Early diagnosis of stroke type is essential before any treatment can be provided. In IS, antiplatelet, thrombolysis, and thrombectomy form the cornerstone of treatment, whereas, for HS, blood pressure reduction, surgery, embolization techniques, and hemostatic treatment can prevent further damage. The accurate identification of the type of stroke is therefore essential for a timely treatment and improved outcomes.

Unfortunately, there is no single biomarker or even a combination of multiple biomarkers, which differentiate IS from HS. Neuroimaging techniques like Magnetic Resonance Imaging (MRI), Computed Tomography (CT) still remain the gold standard for accurately identifying IS, HS and stroke mimics. Due to the high cost associated, neuroimaging facilities are mostly limited to major healthcare institutions and laboratories. The availability becomes even more scarce in sub-urban and rural areas of developing countries.

Clinical scores to identify and classify strokes are an affordable technique which can be made widely accessible for use by primary healthcare workers, lab technicians, and paramedics. Clinical Scores for SC are a weighted linear combination of various clinical attributes that are statistically significant for the identification of type of stroke. Such tools capable of satisfactory performance metrics can make a difference in managing stroke in the resource limited settings prevailing in developing countries. Though such a score may not replace neuroimaging for identifying the type of stroke, it can hasten initiation of stroke management. Scores that can identify strokes and further classify it as IS within a window period, prior to neuroimaging, may prompt immediate referral to a stroke ready hospital for intervention. Management of blood pressure before referral for neuroimaging may also be guided by a reliable and accurate clinical score.

In this paper, we reanalysed the validation studies on clinical scores for SC, from January 2001 to April 2021, to demonstrate the potential of a simple linear combination of clinical attributes for the accurate diagnosis IS and HS. We analysed studies from different demographics, that have validated the results of four popular stroke scores, namely Siriraj Score (SS), Guys Hospital Score/Allen Score (GHS/AS), Greek Score (GS), Besson Score (BS) [5–8]. SSS, GHS/AS, and GS have been designed to classify CT verified cases of IS and HS into three categories of IS, HS and Equivocal (EQ). Whereas BS aims to differentiate IS from the rest. A glossary of machine learning terminologies used in this paper are present in Table 1. Table 2 contains a detailed comparison of four scores.

**Table 1:**
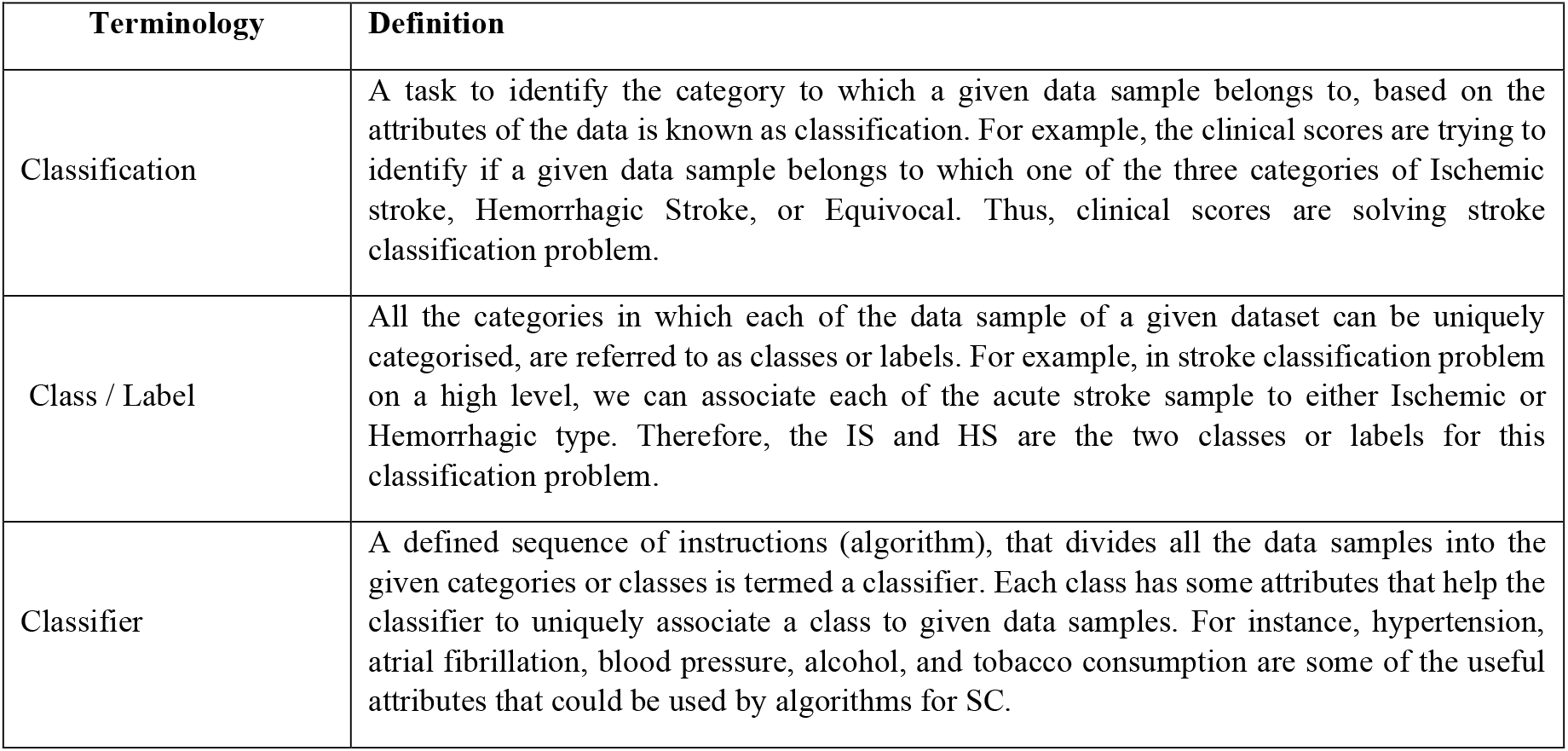
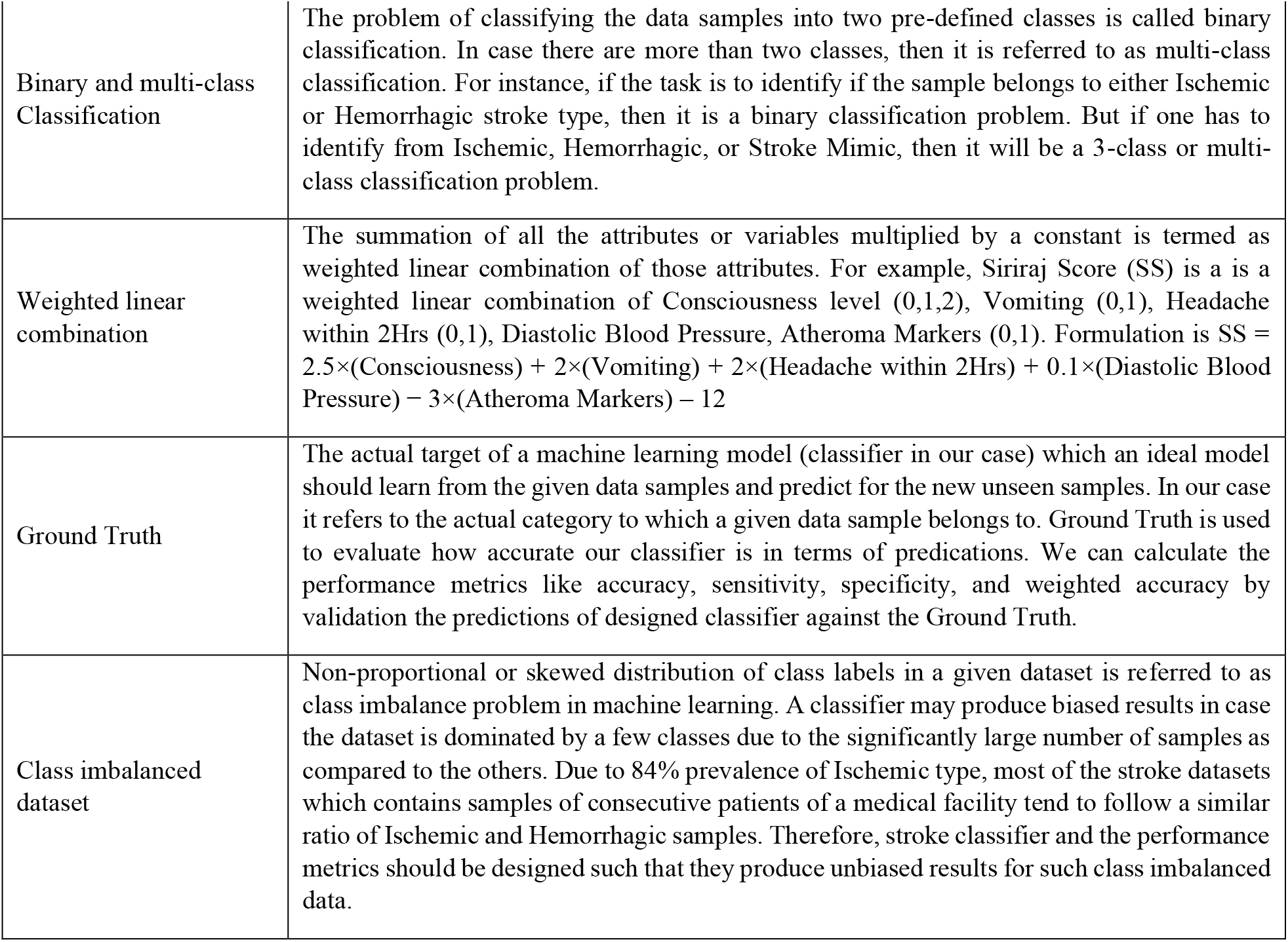
Glossary of machine learning and statistical terminology used in the study.

**Table 2:**
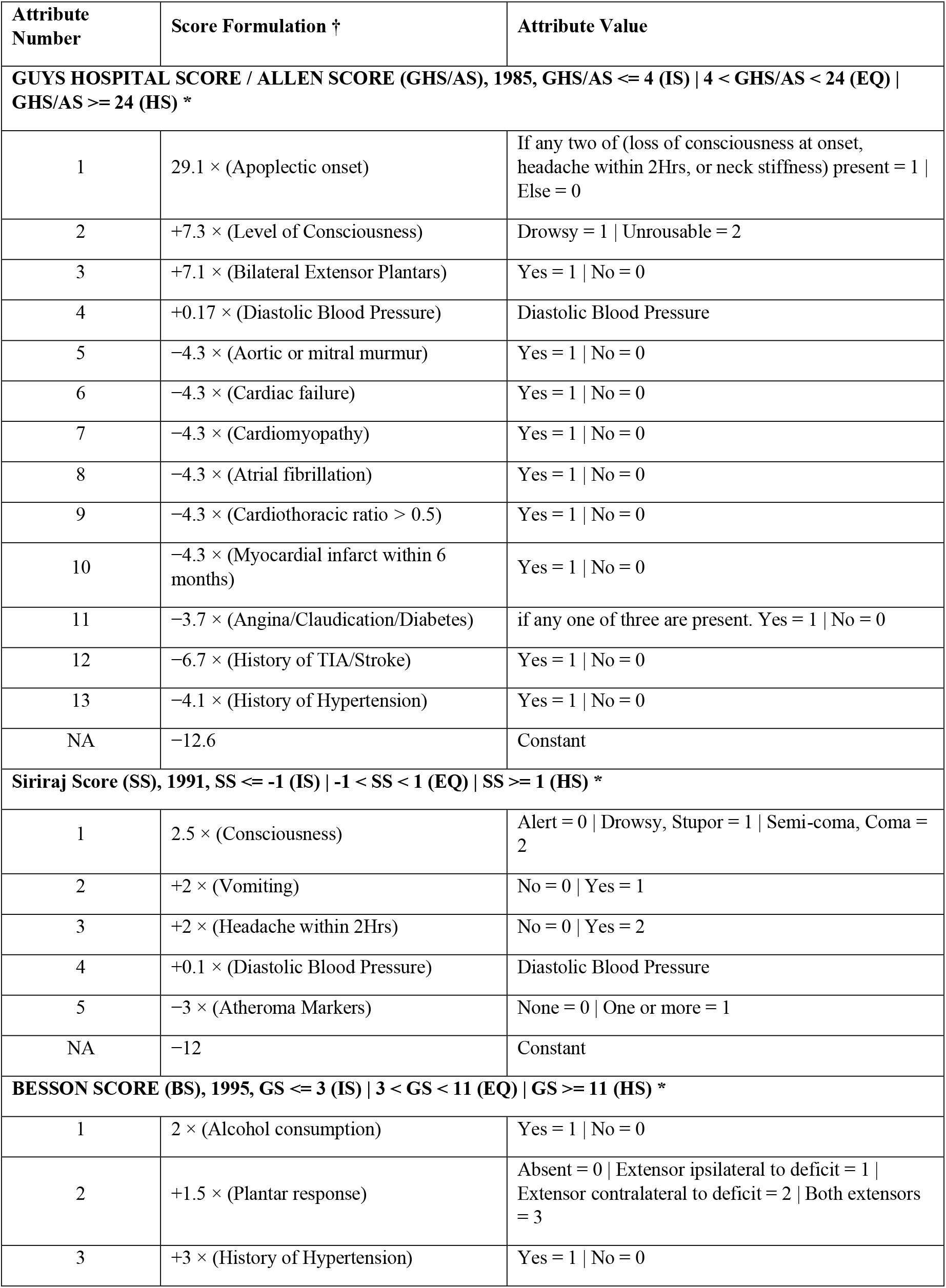
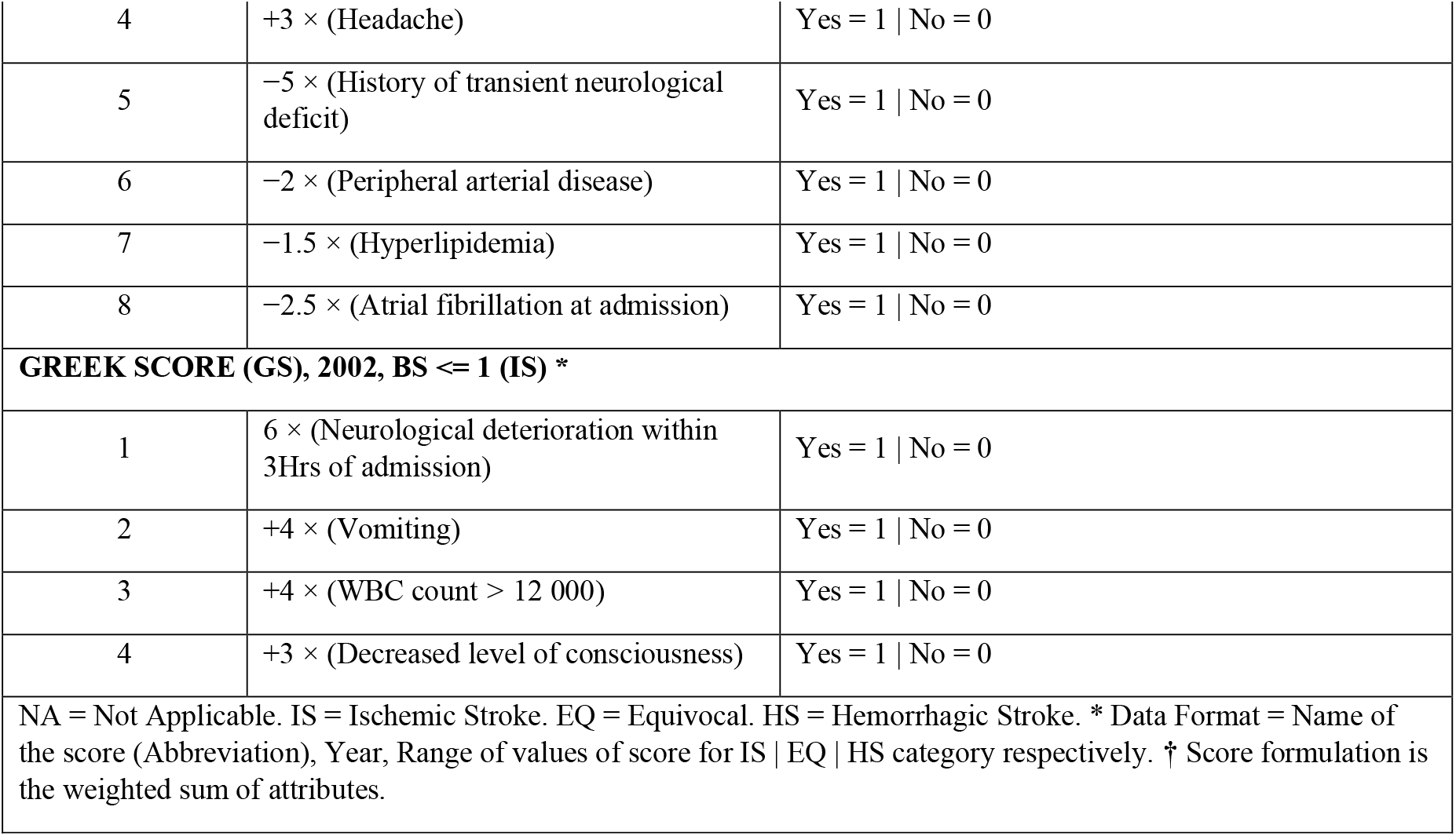
Summary of clinical scores with range of values for ischemic, hemorrhagic and equivocal category predictions.

## Methods

### Search strategy and selection criteria

We performed an advanced search on bibliographic databases PubMed, ScienceDirect, IEEE Xplore, ERIC from January 2001 to April 2021. Also, a comprehensive literature search was done to identify the related studies. Search queries included keywords (Siriraj, Allen, Guys Hospital, Greek, Besson) Score, Stroke Classification, Cerebral (Ischemia, Hemorrhage), Clinical Stroke Identification, Clinical Stroke Score and Stroke in Resource Limited Setting.

Studies that evaluated the stroke classification performance of SS, GHS/AS, GS and BS against the CT scan results as the ground truth have been included in the study. The studies which perform SC using neuroimaging as any of the attributes have been excluded. The other exclusion criteria were studies before 2001, ambiguity in data reported, same data reported in two studies by the same author, studies published in a language other than English, and the studies that have reported results for only one of the two diagnosis of either IS or HS.

The duplicates were removed based on title, first author, and year of publication. Abstracts of 464 uniquely identified studies were then analysed by a reviewer to exclude non-relevant studies. Fig 1 PRISMA flow diagram shows the step-by-step methodology for study selection and the number of studies excluded at each step. A total of 21 studies were finally included. The summary of all the studies has been provided in Table 3.

**Table 3:** Summary of the 21 studies included in the Article.

| Study Name | Scores Validated | Converted SC to two binary classification problem? | Included EQ in performance | Did not encounter EQ |
| --- | --- | --- | --- | --- |
| Boke '21 (India) [13] | 1 (SS) | No | No | No |
| Ravi '20 (India) [14] | 1 (SS) | No | No | No |
| Chakraborty '17 (India) [15] | 1 (SS) | No | No | No |
| Chaudhari '17 (India) [16] | 2 (SS, GHS/AS) | No | Yes (SS) | Yes (GHS/AS) |
| Somasundaran '17 (India) [17] | 1 (SS) | No | No | No |
| Goswami '13 (India) [18] | 4 (SS, GHS/AS, GS, BS) | Yes (SS, GHS/AS, GS) | Yes (SS, GHS/AS, GS) | Yes (BS) |
| Raghuram '12 (India) [19] | 2 (SS, GHS/AS) | No | No | No |
| Sherin '11 (Pakistan) [11] | 2 (SS, GHS/AS) | Yes (SS, GHS/AS) | Yes (SS, GHS/AS) | No |
| Walker '11 (Tanzania) [12] | 2 (SS, GHS/AS) | No | No | No |
| Nouria '09 (Tunisia) [20] | 2 (SS, GHS/AS) | No | No | No |
| Salawu '09 (Nigeria) [21] | 2 (SS, GHS/AS) | No | No | Yes (SS, GHS/AS) |
| Nyandaiti'08 (Nigeria) [22] | 1 (SS) | No | No | No |
| Berhe '08 (Ethiopia) [23] | 1 (GS) | No | No | No |
| Connor '07 (South Africa) [24] | 2 (SS, GHS/AS) | Yes (SS, GHS/AS) | Yes (SS, GHS/AS) | No |
| Ozeren '06 (Turkey) [25] | 2 (SS, GHS/AS) | No | No | No |
| Kolapo '06 (Nigeria) [26] | 1 (SS) | No | No | No |
| Zenebe '05 (Ethiopia) [27] | 1 (SS) | No | No | No |
| Soman '04 (India) [28] | 3 (SS, GHS/AS, GS) | No | No | No |
| Badam '03 (India) [29] | 2 (SS, GHS/AS) | No | No | No |
| Ogun '02 (Nigeria) [10] | 1 (SS) | No | Yes (SS) | No |
| Wadhwani '02 (India) [30] | 2 (SS, GHS/AS) | No | No | Yes (GHS/AS) |
| Total = 21 | 36 | Yes = 7, No = 29 | Yes = 9, No = 27 | Yes = 5, No = 31 |

**Fig 1.**
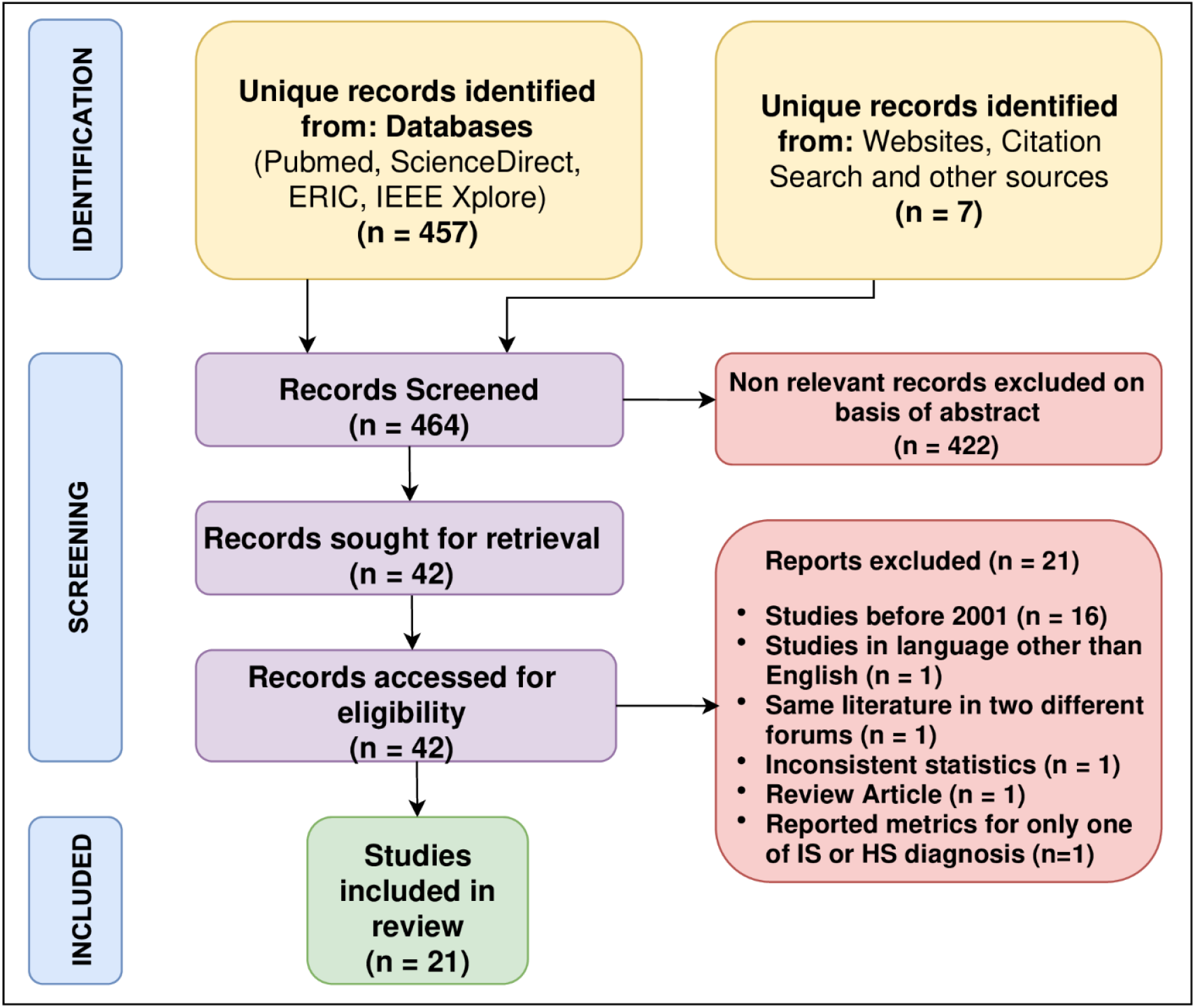
PRISMA flow diagram for study selection.

The email addresses of the authors to make data sharing requests were retrieved from the publications. In case of unavailability of correspondence email in the publication, an internet search was done to reach out to them. We could not the retrieve email addresses of the authors of the four studies. Data sharing requests were made via email to all the authors. Only three authors responded to the request [10–12]. A reminder was further sent to the authors who did not respond.

### Multi-class classification

The task of identifying the category to which a given data sample belongs to, based on the attributes of the data, is known as classification. All the categories in which each of the data sample of a given dataset can be uniquely categorised, are referred to as classes or labels. The problem of dividing the samples into two pre-defined classes is called binary classification. In case there are more than two classes, then it is referred to as multi-class classification. For example, if in SC, the data samples must be categorized in IS or HS classes, then it is a problem of binary classification. But if the categories are IS, HS, and stroke mimics, then it becomes a multi-class classification problem.

A defined sequence of instructions (algorithm), that divides all the data samples into the given categories or classes is termed a classifier. Each class has some attributes that help the classifier to uniquely associate a class to given data samples. For instance, hypertension, atrial fibrillation, blood pressure, alcohol, and tobacco consumption are some of the useful attributes that could be used by algorithms for SC. The performance of a classifier can be measured by widely used evaluation metrics derived from confusion matrix namely, accuracy, sensitivity/specificity, and their combination by geometric mean (GM).

#### Confusion matrix (C)

A cross tabulation that provides a summary of the number of correct and incorrect predictions by a classifier for each of the defined classes. For a N-class classifier, it is a N × N square matrix. The *i*^th^ row contains the samples predicted to be of *i*^th^ class. The diagonal elements represent the number of correctly classified samples of each class. The (*i, j*)^th^ element of the matrix (C_ij_) refers to the number of samples predicted to be from *i*^th^ class, but actually belong to *j*^th^ class. Thus, the number of correct predictions for the *i*^th^ class is the diagonal element (C_ii_) of matrix C. Fig 2 is the representation of 3-class confusion matrix. Where, total samples in class A, B, C respectively = N_A_, N_B_, N_C_, total predictions in class A, B, C respectively = P_A_, P_B_, P_C_, total samples (N) = N_A_ + N_B_ + N_C_ = P_A_ + P_B_ + P_C._

**Fig 2.**
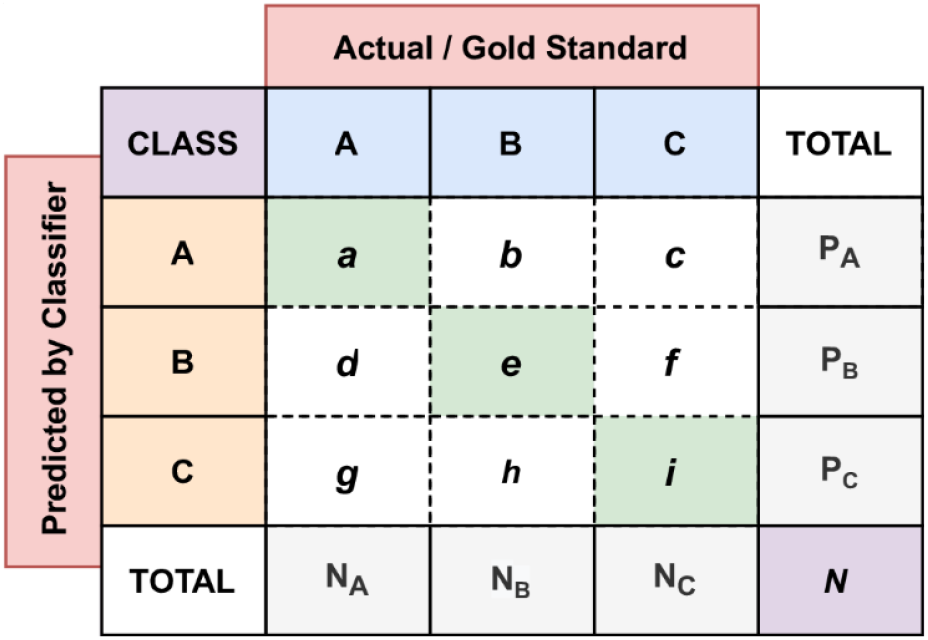
Confusion Matrix (C) for 3-class classification.

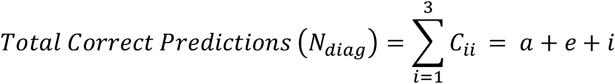

Following metrics have been defined with respect to confusion matrix in Fig 2.

#### Accuracy

It the most popular metric used across domains. It represents the ratio of total number of correctly classified samples of all the classes to total number of samples.

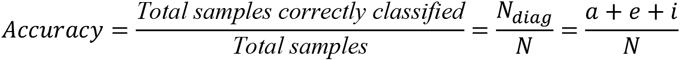

#### Sensitivity or True Positive Rate (TPR) and Specificity or True Negative Rate (TNR)

*Sensitivity or TPR* of a particular class refers to the ability of a classifier to correctly classify that class. Whereas *Specificity or TNR* of the same class, refers to the ability of a classifier to correctly classify the rest of the classes. Considering *i*^th^ class as the positive class, rest all the classes will be the negative classes. Therefore, the total samples that actually belong to class *i* as per the ground truth are called positives (POS) = N_i_ and the sample that actually belong to classes other than class *i* are called negatives (NEG) = N − N_i_. The number of positive class samples correctly predicted as positive are known as true positives (TP), and the samples from negative classes being correctly predicted by the classifier are known as true negatives (TN). Similarly, the samples of positive class wrongly classified as negative are called false negatives (FN) and the samples of negative class wrongly classified as positive are called false positives (FP).

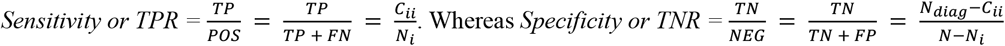

#### Geometric Mean (GM)

GM is the combination of sensitivity and specificity. It is the square root of the product of sensitivity and specificity. 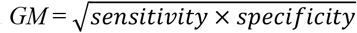

### Class Imbalance Problem

Non-proportional or skewed distribution of class labels in a given dataset is referred to as class imbalance problem in machine learning. A classifier may produce biased results in case the dataset is dominated by a few classes as compared to the others. Most widely used evaluation metrics of accuracy, can be misleading when used for assessing performance of a classifier deployed for class imbalanced cases [31].

Consider a binary classification problem with 100 samples, of which 85 belong to class 1 and the remaining 15 belong to class 2. A trivial classifier that classifies all the samples to class 1, gives the model an accuracy of 85%. The classifier while unable to predict class 2 correctly at all, may appear to perform well in terms of performance using the accuracy metric. Accuracy can be misleading if used for class imbalanced cases.

Weighted Accuracy (WA) is one of the known solutions to the problem of class imbalance, which provides equal importance to all the classes present in the dataset. In a classification problem on a data with *N* samples and *C* classes, the number of samples be *N*_*i*_ and the true positive predictions by the classifier be *TP*_*i*_ for the *i*^*th*^ class. Then the weight of *i*^*th*^ class is defined as 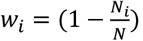, and the weighted accuracy can be mathematically formulated as:

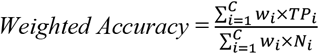

Let’s reconsider the previously cited example of biased binary. For class 1 and 2 respectively, true positives will be *TP*_1_ = 85, *TP*_2_ = 0 and the class weights will be *w*_1_ = 0.15, *w*_2_ = 0.85. The weighted accuracy of the 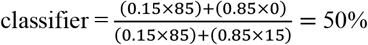. WA clearly shows that the classifier is no better than a random guess on the data.

### Redefining stroke classification as a multi-classification problem

SC, though originally perceived as a binary classification problem (either IS or HS), the clinical scores SS, GHS/AS, and GS introduced a third class labelled as equivocal/indeterminate (EQ). Introduction of the new class by the classifiers, modifies SC to a 3-class classification problem. It can be considered as the number of samples for the third class *N*_*EQ*_ are *zero* as per the gold standard. It implies that the samples which are classified as EQ, actually belong to either of IS or HS class. Therefore, EQ will always contribute to incorrectly classified samples by the classifier, thus reducing the performance. Excluding them during the evaluation, as done by most of the analysed studies, results in higher performance metric values than actual. Hence, we argue that they should be included while evaluating the models to get the accurate estimate of the performance.

### Redefining stroke classification as a class imbalance problem

SC needs to be approached as a class imbalance problem as the prevalence of IS in the dataset is approximately 84% [2]. Therefore, using accuracy as one of the performance metrics may lead to over optimistic estimates of the classifier performance.

In the next section we re-evaluate the results of all the studies using WA measure and compare with the corresponding accuracy measures by viewing SC as a problem characterised by multi-class classification and class imbalance.

## Results

We extracted data for a total of 36 evaluation of scores from 21 studies. The details have been provided in Table 2. SS has been evaluated by 20 studies, GHS/AS by 12, GS by three, and BS by only one study. Out of 36 evaluations, seven converted SC problem to two separate binary classification problems with IS/Non-IS classes and HS/Non-HS classes [11,18,24]. They reported a separate confusion matrix for each of the two classifiers. For IS diagnosis, they include EQ cases in Non-IS class and in Non-HS class during HS diagnosis. Such a practice is not correct for a binary classification problem. The rest of the studies classified SC as IS versus HS. Ideally for a binary classification problem such that of SC, the sensitivity of IS diagnosis should be equal to specificity of HS diagnosis and vice-versa. But for the studies that have converted the problem to two separate classification problems, this property will not be applicable. Therefore, to maintain consistency we have reported sensitivity of IS and HS instead of sensitivity and specificity of the classifier during our further analysis.

Only nine evaluations considered EQ for the calculation of performance metrics. Five evaluations did not encounter EQ cases, and the rest excluded EQ predictions performance evaluation leading to the over-estimates of performance results.

The range, aggregate, and median of the reported and calculate metrics for all scores have been in Table 4 which is based on the analysis done on the data extracted from the included studies. For the details of sample size, percentage class distribution, and confusion matrix (cross tabulation) reported by the studies reader can refer to Table 5. Based on these reported numbers, we created a confusion matrix including EQ cases for IS diagnosis for each study in Table 6. The sensitivity and specificity reported by all the studies and calculated performance metrics from the corrected confusion matrix have been tabulated in Table 6. The confusion matrix for HS Diagnosis can be retrieved by interchanging of table columns, TP (True Positive) with TN (True Negative), and FP (False Positive) with FN (False Negative).

**Table 4:** Range, aggregate and median of reported and calculated sensitivity for IS diagnosis and HS diagnosis, reported and calculated accuracy, and calculated weighted accuracy.

| Evaluation Metric | Range |  |  |  | Aggregate / Mean |  |  |  | Median |  |  |  |
| --- | --- | --- | --- | --- | --- | --- | --- | --- | --- | --- | --- | --- |
|  | SS | GHS/AS | GS | BS | SS | GHS/AS | GS | BS | SS | GHS/AS | GS | BS |
| Reported Sensitivity (IS) | 43-97% | 35-94% | 74-94% | 66% | NA<sup>†</sup> | NA<sup>†</sup> | NA<sup>†</sup> | NA<sup>†</sup> | 78% (65-88%) | 73% (62-88%) | 89% (81-92%) | 66% |
| Reported Sensitivity (HS) | 50-95% | 33-93% | 41-80% | NA | NA<sup>†</sup> | NA<sup>†</sup> | NA<sup>†</sup> | NA<sup>†</sup> | 71% (64-82%) | 63% (39-73%) | 78% (59-79%) | NA* |
| Calculated Sensitivity (IS) | 40-90% | 35-93% | 60-74% | 66% | 71% (15%) | 64% (17%) | 69% (7%) | 66% (0%) | 70% (57-73%) | 64% (53-71%) | 64% (62-69%) | 66% |
| Calculated Sensitivity (HS) | 34-86% | 20-73% | 11-80% | NA | 61% (17%) | 44% (17%) | 51% (35%) | NA* | 59% (50-79%) | 46% (34-64%) | 43% (27-62%) | NA* |
| Reported Accuracy | 47-91% | 40-87% | 84-84%<br>‡ | NA* | NA<sup>†</sup> | NA<sup>†</sup> | NA<sup>†</sup> | NA<sup>†</sup> | 80% (59-85%) | 56% (48-72%) | 84% (84-84%)<br>‡ | NA* |
| Calculated Accuracy | 37-86% | 40-87% | 38-76% | NA* | 68% (13%) | 59% (14%) | 61% (19%) | NA* | 67% (56-75%) | 58% (47-61%) | 51% (45-63%) | NA* |
| Calculated Weighted Accuracy | 37-85% | 43-80% | 38-77% | NA* | 66% (12%) | 54% (13%) | 60% (20%) | NA* | 64% (54-73%) | 53% (47-62%) | 51% (44-64%) | NA* |
| Data format of Aggregate Column = Mean (SD). Data format of Median Column = Median (IQR). All values have been rounded up to nearest integer value. NA* = Values not applicable for BS as it reports only IS diagnosis metrics. NA<sup>†</sup> = Aggregate values could not be computed as some of the studies include EQ while the rest exclude them. ‡ = Value reported by only one study. Therefore, range is not applicable. |  |  |  |  |  |  |  |  |  |  |  |  |

**Table 5:**
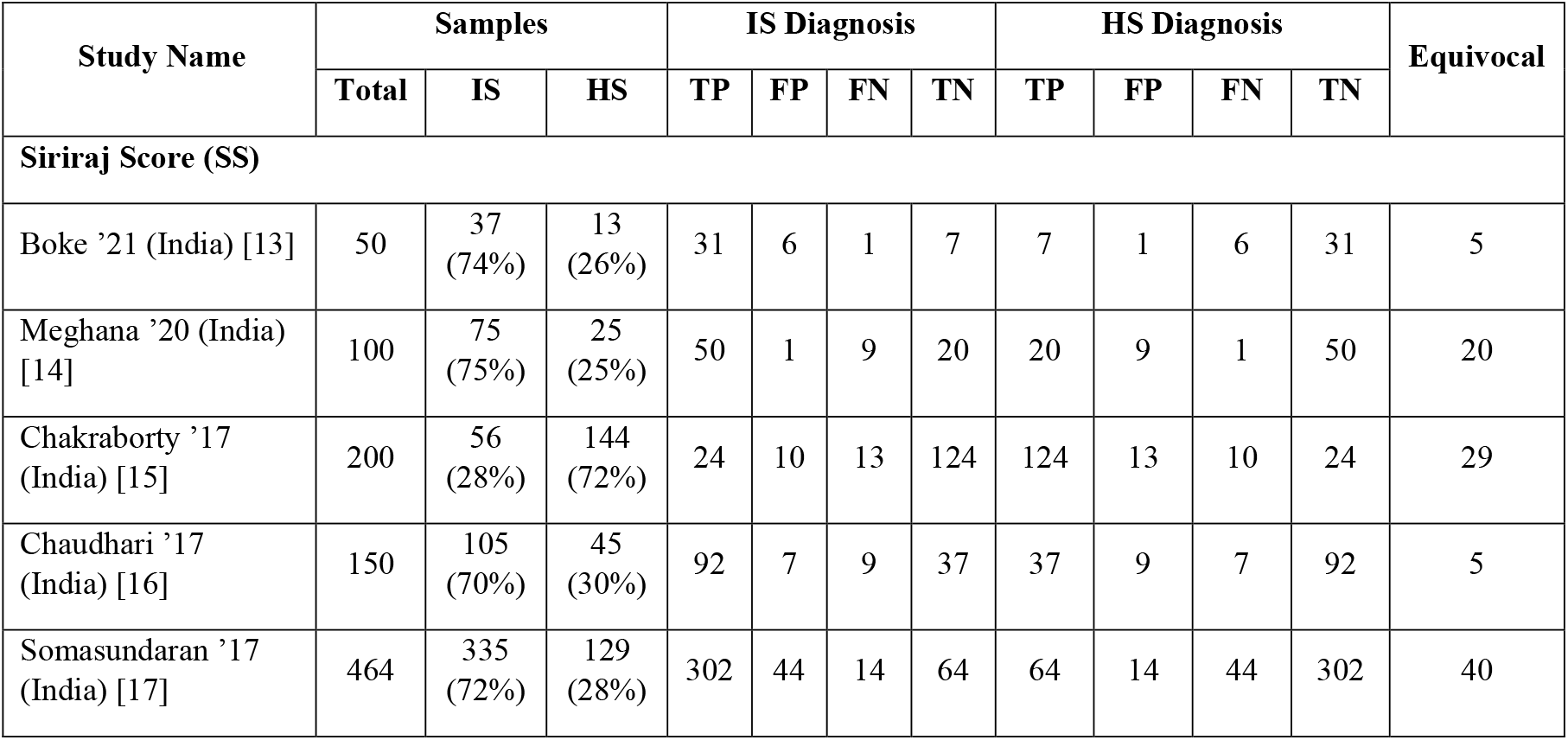
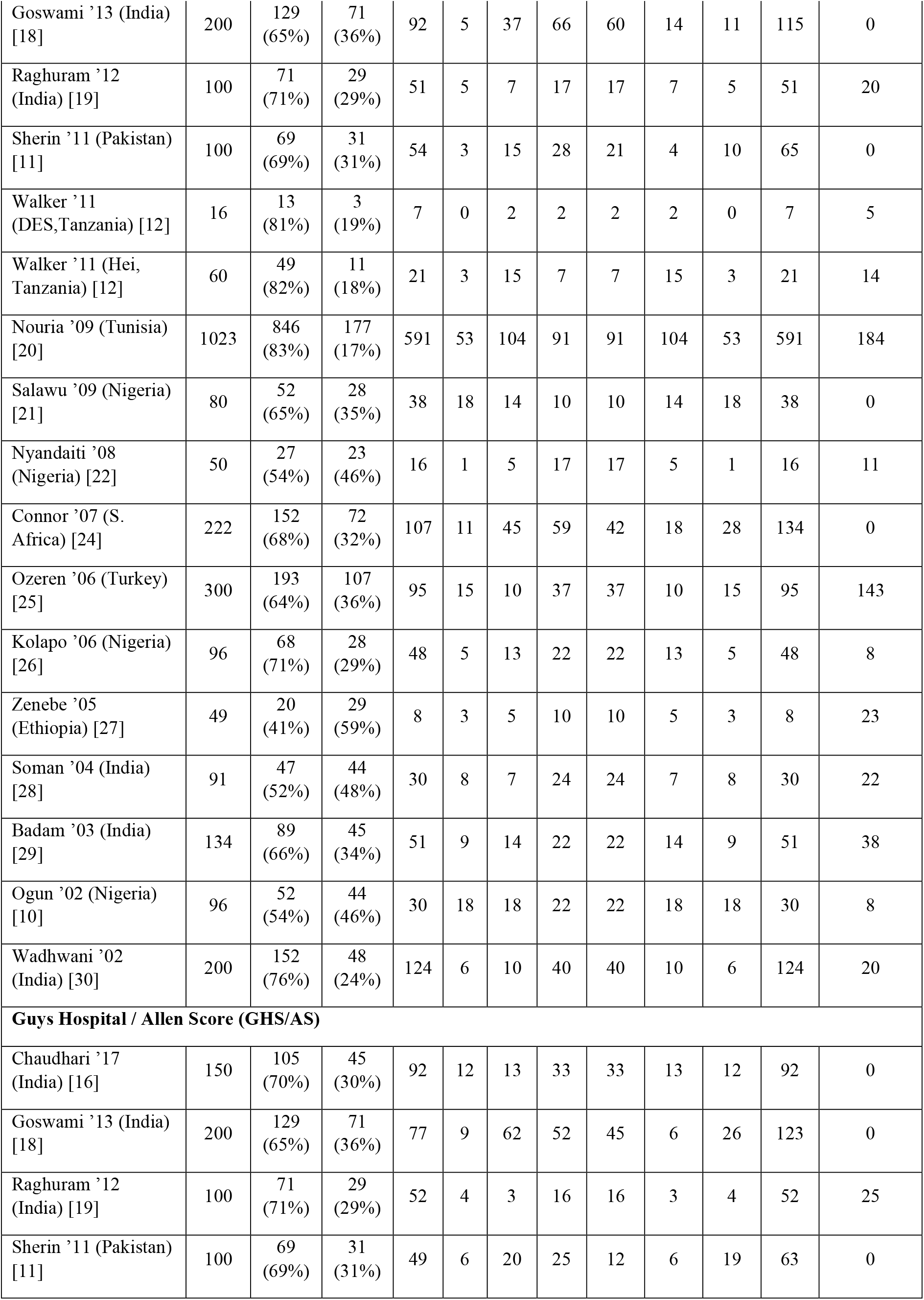
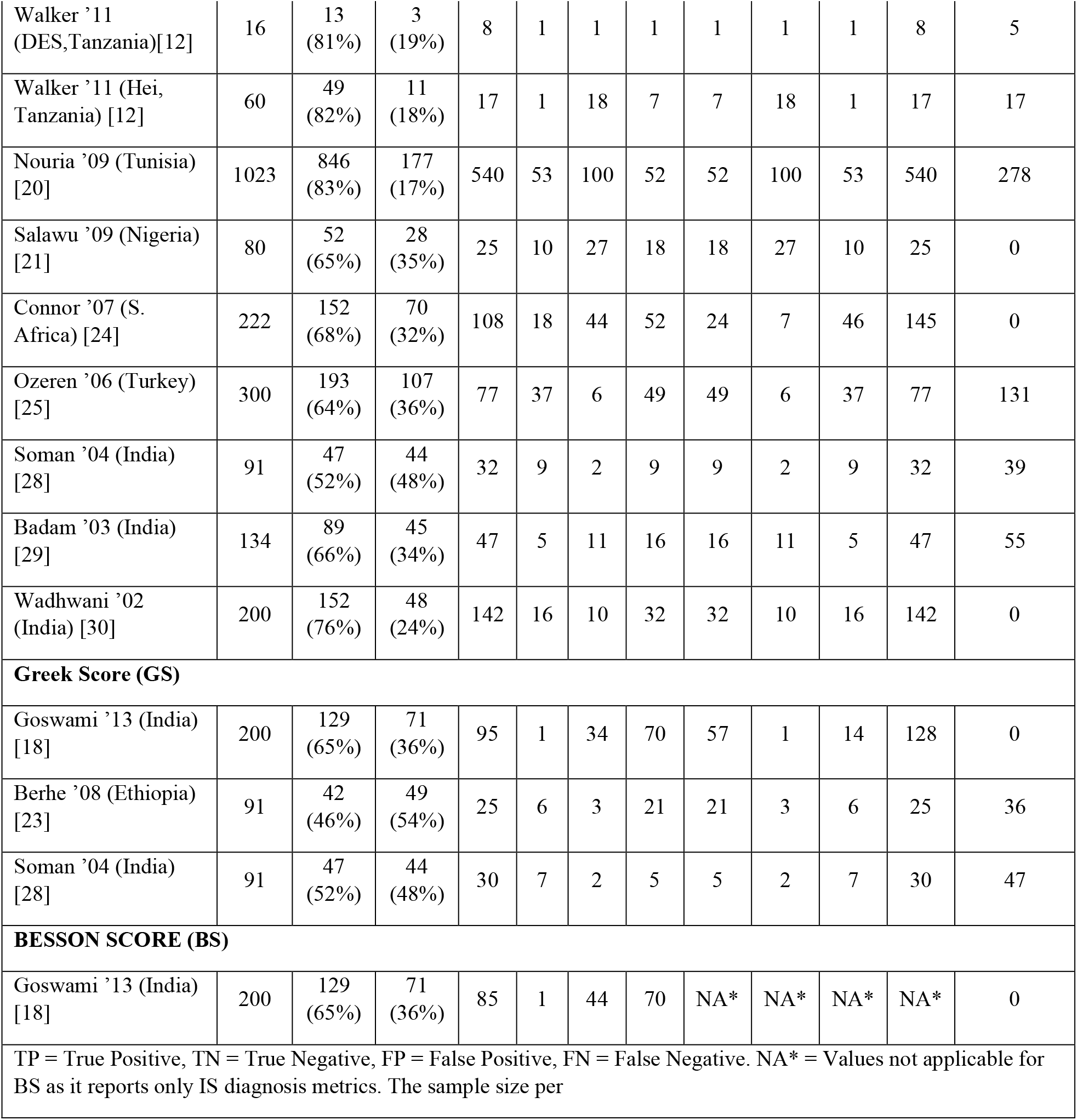
Reported cross tabulation (confusion matrix) for stroke classification against ground truth CT.

**Table 6:**
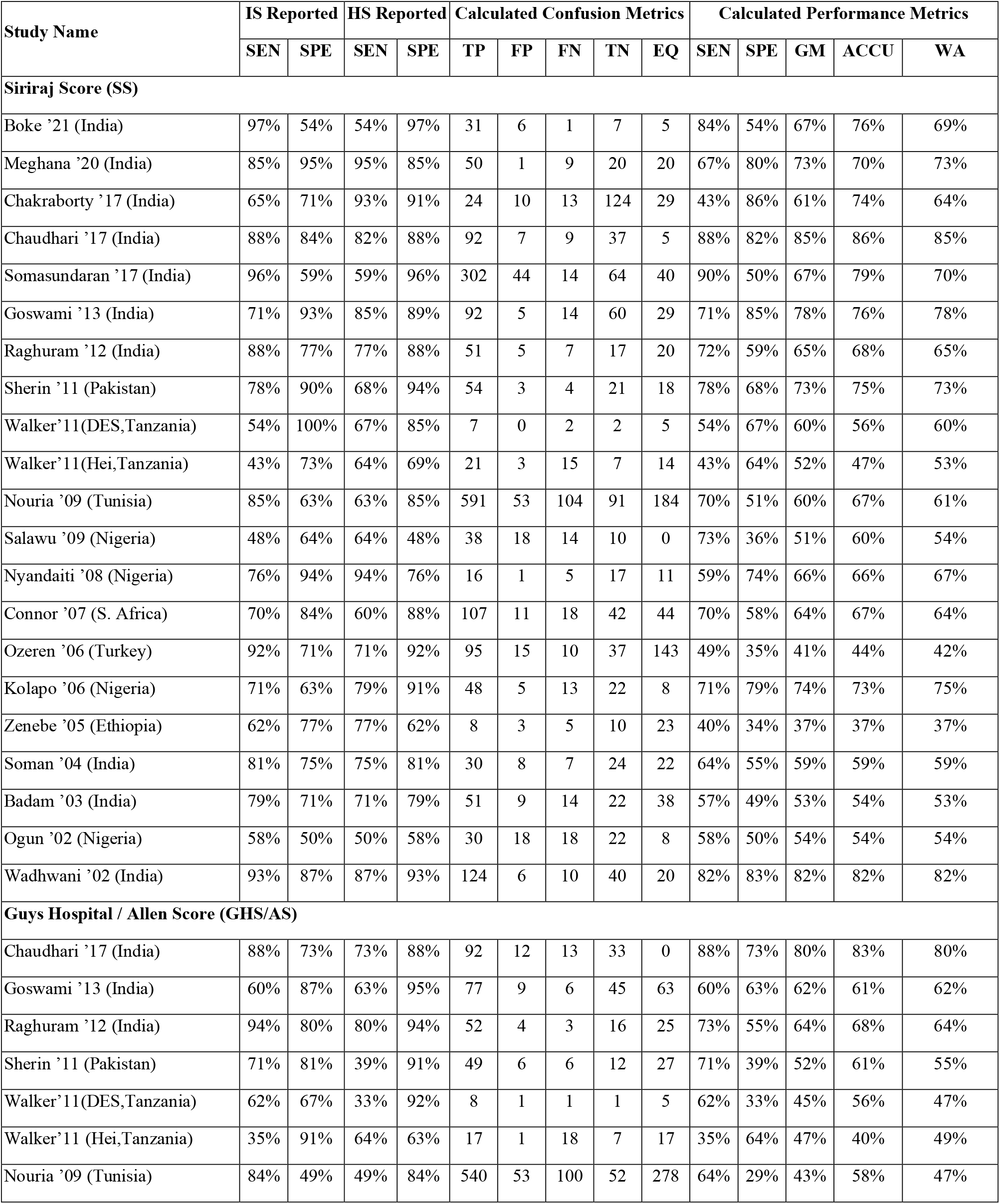
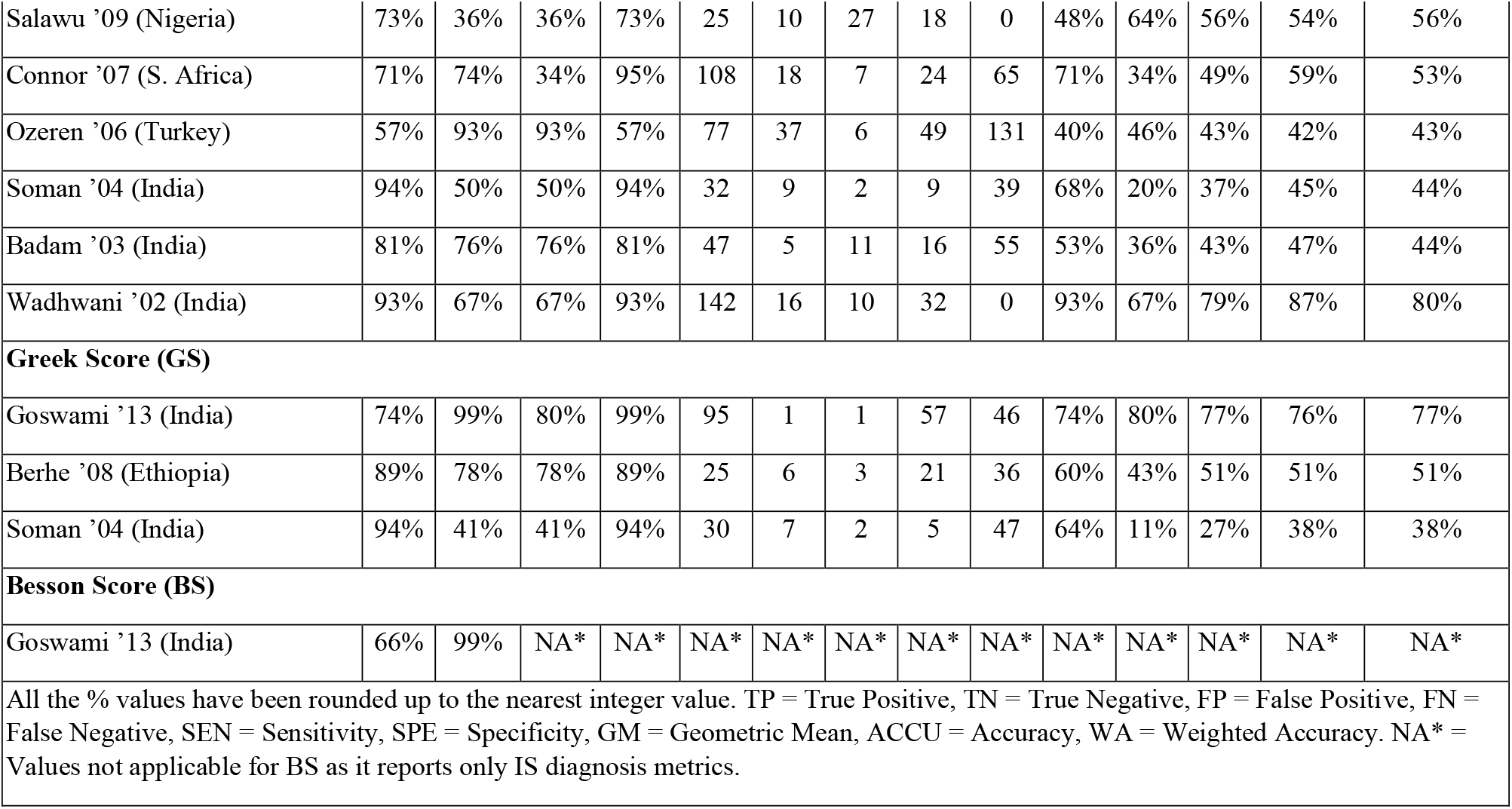
Reported performance metrics by the studies, and corresponding calculated performance metrics based on the corrected confusion metrics (cross tabulation) including equivocal cases.

| Study Name | IS Reported |  | HS Reported |  | Calculated Confusion Metrics |  |  |  |  | Calculated Performance Metrics |  |  |  |  |
| --- | --- | --- | --- | --- | --- | --- | --- | --- | --- | --- | --- | --- | --- | --- |
|  | SEN | SPE | SEN | SPE | TP | FP | FN | TN | EQ | SEN | SPE | GM | ACCU | WA |
| **Siriraj Score (SS)** |  |  |  |  |  |  |  |  |  |  |  |  |  |  |
| Boke '21 (India) | 97% | 54% | 54% | 97% | 31 | 6 | 1 | 7 | 5 | 84% | 54% | 67% | 76% | 69% |
| Meghana '20 (India) | 85% | 95% | 95% | 85% | 50 | 1 | 9 | 20 | 20 | 67% | 80% | 73% | 70% | 73% |
| Chakraborty '17 (India) | 65% | 71% | 93% | 91% | 24 | 10 | 13 | 124 | 29 | 43% | 86% | 61% | 74% | 64% |
| Chaudhari '17 (India) | 88% | 84% | 82% | 88% | 92 | 7 | 9 | 37 | 5 | 88% | 82% | 85% | 86% | 85% |
| Somasundaran '17 (India) | 96% | 59% | 59% | 96% | 302 | 44 | 14 | 64 | 40 | 90% | 50% | 67% | 79% | 70% |
| Goswami '13 (India) | 71% | 93% | 85% | 89% | 92 | 5 | 14 | 60 | 29 | 71% | 85% | 78% | 76% | 78% |
| Raghuram '12 (India) | 88% | 77% | 77% | 88% | 51 | 5 | 7 | 17 | 20 | 72% | 59% | 65% | 68% | 65% |
| Sherin '11 (Pakistan) | 78% | 90% | 68% | 94% | 54 | 3 | 4 | 21 | 18 | 78% | 68% | 73% | 75% | 73% |
| Walker'11(DES,Tanzania) | 54% | 100% | 67% | 85% | 7 | 0 | 2 | 2 | 5 | 54% | 67% | 60% | 56% | 60% |
| Walker'11(Hei,Tanzania) | 43% | 73% | 64% | 69% | 21 | 3 | 15 | 7 | 14 | 43% | 64% | 52% | 47% | 53% |
| Nouria '09 (Tunisia) | 85% | 63% | 63% | 85% | 591 | 53 | 104 | 91 | 184 | 70% | 51% | 60% | 67% | 61% |
| Salawu '09 (Nigeria) | 48% | 64% | 64% | 48% | 38 | 18 | 14 | 10 | 0 | 73% | 36% | 51% | 60% | 54% |
| Nyandaiti '08 (Nigeria) | 76% | 94% | 94% | 76% | 16 | 1 | 5 | 17 | 11 | 59% | 74% | 66% | 66% | 67% |
| Connor '07 (S. Africa) | 70% | 84% | 60% | 88% | 107 | 11 | 18 | 42 | 44 | 70% | 58% | 64% | 67% | 64% |
| Ozeren '06 (Turkey) | 92% | 71% | 71% | 92% | 95 | 15 | 10 | 37 | 143 | 49% | 35% | 41% | 44% | 42% |
| Kolapo '06 (Nigeria) | 71% | 63% | 79% | 91% | 48 | 5 | 13 | 22 | 8 | 71% | 79% | 74% | 73% | 75% |
| Zenebe '05 (Ethiopia) | 62% | 77% | 77% | 62% | 8 | 3 | 5 | 10 | 23 | 40% | 34% | 37% | 37% | 37% |
| Soman '04 (India) | 81% | 75% | 75% | 81% | 30 | 8 | 7 | 24 | 22 | 64% | 55% | 59% | 59% | 59% |
| Badam '03 (India) | 79% | 71% | 71% | 79% | 51 | 9 | 14 | 22 | 38 | 57% | 49% | 53% | 54% | 53% |
| Ogun '02 (Nigeria) | 58% | 50% | 50% | 58% | 30 | 18 | 18 | 22 | 8 | 58% | 50% | 54% | 54% | 54% |
| Wadhwani '02 (India) | 93% | 87% | 87% | 93% | 124 | 6 | 10 | 40 | 20 | 82% | 83% | 82% | 82% | 82% |
| **Guys Hospital / Allen Score (GHS/AS)** |  |  |  |  |  |  |  |  |  |  |  |  |  |  |
| Chaudhari '17 (India) | 88% | 73% | 73% | 88% | 92 | 12 | 13 | 33 | 0 | 88% | 73% | 80% | 83% | 80% |
| Goswami '13 (India) | 60% | 87% | 63% | 95% | 77 | 9 | 6 | 45 | 63 | 60% | 63% | 62% | 61% | 62% |
| Raghuram '12 (India) | 94% | 80% | 80% | 94% | 52 | 4 | 3 | 16 | 25 | 73% | 55% | 64% | 68% | 64% |
| Sherin '11 (Pakistan) | 71% | 81% | 39% | 91% | 49 | 6 | 6 | 12 | 27 | 71% | 39% | 52% | 61% | 55% |
| Walker'11(DES,Tanzania) | 62% | 67% | 33% | 92% | 8 | 1 | 1 | 1 | 5 | 62% | 33% | 45% | 56% | 47% |
| Walker'11 (Hei,Tanzania) | 35% | 91% | 64% | 63% | 17 | 1 | 18 | 7 | 17 | 35% | 64% | 47% | 40% | 49% |
| Nouria '09 (Tunisia) | 84% | 49% | 49% | 84% | 540 | 53 | 100 | 52 | 278 | 64% | 29% | 43% | 58% | 47% |
| Salawu '09 (Nigeria) | 73% | 36% | 36% | 73% | 25 | 10 | 27 | 18 | 0 | 48% | 64% | 56% | 54% | 56% |
| Connor '07 (S. Africa) | 71% | 74% | 34% | 95% | 108 | 18 | 7 | 24 | 65 | 71% | 34% | 49% | 59% | 53% |
| Ozeren '06 (Turkey) | 57% | 93% | 93% | 57% | 77 | 37 | 6 | 49 | 131 | 40% | 46% | 43% | 42% | 43% |
| Soman '04 (India) | 94% | 50% | 50% | 94% | 32 | 9 | 2 | 9 | 39 | 68% | 20% | 37% | 45% | 44% |
| Badam '03 (India) | 81% | 76% | 76% | 81% | 47 | 5 | 11 | 16 | 55 | 53% | 36% | 43% | 47% | 44% |
| Wadhwani '02 (India) | 93% | 67% | 67% | 93% | 142 | 16 | 10 | 32 | 0 | 93% | 67% | 79% | 87% | 80% |
| **Greek Score (GS)** |  |  |  |  |  |  |  |  |  |  |  |  |  |  |
| Goswami '13 (India) | 74% | 99% | 80% | 99% | 95 | 1 | 1 | 57 | 46 | 74% | 80% | 77% | 76% | 77% |
| Berhe '08 (Ethiopia) | 89% | 78% | 78% | 89% | 25 | 6 | 3 | 21 | 36 | 60% | 43% | 51% | 51% | 51% |
| Soman '04 (India) | 94% | 41% | 41% | 94% | 30 | 7 | 2 | 5 | 47 | 64% | 11% | 27% | 38% | 38% |
| **Besson Score (BS)** |  |  |  |  |  |  |  |  |  |  |  |  |  |  |
| Goswami '13 (India) | 66% | 99% | NA* | NA* | NA* | NA* | NA* | NA* | NA* | NA* | NA* | NA* | NA* | NA* |
| All the % values have been rounded up to the nearest integer value. TP = True Positive, TN = True Negative, FP = False Positive, FN = False Negative, SEN = Sensitivity, SPE = Specificity, GM = Geometric Mean, ACCU = Accuracy, WA = Weighted Accuracy. NA* = Values not applicable for BS as it reports only IS diagnosis metrics. |  |  |  |  |  |  |  |  |  |  |  |  |  |  |

For SS, the reported sensitivity for IS diagnosis was 43-97% (median 78% [IQR 65-88%]) and for HS diagnosis it ranged from 50-95% (median 71% [IQR 64-82%]). The corresponding calculated values of the sensitivity for IS diagnosis was 40-90% (median 70% [IQR 57-73%], aggregate 71% [SD 15%]) and for HS diagnosis it ranged from 34-86% (median 59% [IQR 50-79%], aggregate 61% [SD 17%]). The reported accuracy was 47-91% (80% [IQR 59-85%]), whereas the calculated accuracy values was 37-86% (median 67% [IQR 56-75%], aggregate 68% [SD 13%]). However, when the class imbalance was considered, the weighted accuracy ranged from 37-85% (median 64% [IQR 54-73%], aggregate 66% [SD 12%]).

For GHS/AS, the reported sensitivity for IS diagnosis was 35-94% (median 73% [IQR 62-88%]) and for HS diagnosis it ranged from 33-93% (median 63% [IQR 39-73%]). The sensitivity for IS diagnosis was 35-93% (median 64% [IQR 53-71%], aggregate 64% [SD 17%]) and for HS diagnosis it was 20-73% (median 46% [IQR 34-64%], aggregate 44% [SD 17%]). The reported accuracy ranged from 40-87% (median 56% [IQR 48-72%]), whereas the calculated accuracy values was 40-87% (median 58% [IQR 47-61%], aggregate 59% [SD 14%]). The weighted accuracy was 43-80% (median 53% [IQR 47-62%], aggregate 54% [SD 13%]).

For GS, the reported sensitivity for IS diagnosis was 73-94% (median 89% [IQR 81-92%]) and for HS diagnosis it ranged from 41-80% (median 78% [IQR 59-79%]). The corresponding calculated values of the sensitivity for IS diagnosis was 60-74% (median 64% [IQR 62-69%], aggregate 69% [SD 7%]) and for HS diagnosis it ranged from 11-80% (median 43% [IQR 27-62%], aggregate 51% [SD 35%]). Only one study reported the accuracy for GS of 84%, whereas the calculated accuracy values ranged from 38-76% (median 51% [IQR 45-63%], aggregate 61% [SD 19%]). The weighted accuracy was 38-77% (median 51% [IQR 44-64%], aggregate 60% [SD 20%]).

As BS performs single class classification of IS/Non-IS, therefore HS diagnosis results and other performance metrics for multi-class classification including EQ cases could not be calculated.

Total number of samples across studies over which the scores have been evaluated are 3781 for SS, 2676 for GHS/AS, 382 for GS and 200 for BS. As per the aggregate and median results in Table 4, SS performed better than the others. On the studies that have compared multiple scores on the same set of patients, we performed *t-test* to count the number of studies in which SS performed better than GHS/AS and GS and vice-versa. As per the results of *t-test*, SS performed better than GHS/AS in eight studies, whereas GHS/AS showed better performance than SS in four studies. Also, SS performed better than GS in 2 studies while GS performed better in none of the studies. In *t-test* of GHS/AS and GS, both the scores are performed better than the other in one-one study each. Also, the median and aggregate results for these two scores were inconsistent.

SS and GHS/AS have been validated by significant number of studies, but GS and BS lack in terms of number of validations. Aggregate and median values of performance metrics showed consensus with *t-test* results for SS. Therefore, we can conclude that it performs better than the other scores. However, due to inconsistency in results of GHS/AS and GS, no conclusion could be made. Moreover, GS and BS require more evaluations globally.

Studies from various demographics showed high variability in the performance of the scores. Therefore, the scores need to be fine-tuned for different geographic locations as they don’t generalize globally. These studies have been conducted at different medical facilities and due to unavailability of public datasets, the results could not be reproduced and validated. Due to the same reason the factors for such a high variability in results have not been analysed and are still unknown.

We reached out to a total of 25 authors of the 21 studies and 4 scores via email with a request to make dataset available. Only 3 responded with a willingness to share the data [10–12]. Lack of publicly available dataset is one of the reasons of poor reproducibility of results in medical researches [32–35].

More than 85% of studies have been conducted on a sample size of less than 200, which is extremely small to get an estimate of generalizability of the scores.

The exclusion of EQ cases during the performance evaluation leads to overestimation of the performance of scores [Table 4]. The reported ranges of metrics were significantly higher than the corresponding calculated value ranges, and their aggregate and median values after including EQ cases.

The ratio of IS and HS samples in the data used in most of the studies, followed a similar trend as that of global prevalence ratio IS to HS, which is heavily skewed towards IS. Relatively higher values of calculated accuracy than WA and their aggregates were noted in SC, which is inherently a case of class imbalance problem.

## Discussion and Conclusion

In this paper, we critically examined SS, GHS/AS, GS and BS for clinical classification of stroke. SC has been defined on a high level as a binary classification problem, but SS, GHS/AS, GS predicted a third class of samples as equivocal. Therefore, we emphasize upon considering SC as a multi-class classification problem for these clinical scores and include EQ cases during evaluation of scores in order to get the correct estimates of the performance. We further explain the need to consider SC as a class imbalance problem due to the non-proportionate prevalence ratio of IS and HS samples in the data. Consequently, we recommend WA instead of accuracy as a measure of performance to avoid over estimation.

SS performed relatively better than the other scores. The results of the aggregate, and median values of metrics, and the *t-test* show consistency for SS when compared with other scores. Any conclusion on the performance of GHA/AS and GS could not be made due to inconsistency in results. Moreover, GS and BS have not been widely evaluated.

To analyse the underlying reasons for high variability in the results of all the scores, we encourage the practice of sharing of anonymized raw data by the authors. It will enhance the reproducibility of the existing research and will support further research in the field.

The performance of SS demonstrates the potential of weighted linear combination of clinical features can prove to be quite significant in accurate identification of stroke type.

We believe that various data driven models based on sophisticated machine learning and deep learning (ML & DL) models would be able to improve the sensitivity, specificity, and weighted accuracy by identifying the latent patterns from the data which we are not able to achieve by a simple linear combination based classifier [36,37]. We further believe that incorporating data on gait and face using computer vision (CV) may further improve the results. ML, DL and CV based techniques may unlock better performance from a classifier.

We recommend data pooling from different demographics for creation of a global clinical dataset with statistically important attributes for stroke classification. It will significantly improve the performance of a classifier while enabling the healthcare worker in resource limited settings, access to early diagnosis of stroke type. A global pooling of raw data will help in generalizing the results for different demographics.

## Data Availability

All data produced in the present work are contained in the manuscript

